# Toward unraveling the correlates of fatigue in glioma

**DOI:** 10.1101/2022.07.29.22277995

**Authors:** Jantine G. Röttgering, Vera Belgers, Philip C. De Witt Hamer, Hans Knoop, Linda Douw, Martin Klein

**Author notes:** Corresponding author, address: Amsterdam UMC location Vrije Universiteit Amsterdam, Medical Psychology, Boelelaan 1117, Amsterdam, The Netherlands. **<u>Funding:</u>** Funding for this work was provided by the Stichting Anita Veldman Foundation (CCA-2019-2-21). **<u>Authorship:</u>** Conceptualization and design: JR, MK, LD. Data collection and curation: JR, VB. Data analysis and interpretation: JR, VB, LD, MK. Visualization: JR. Writing of the manuscript: JR. Revising of the manuscript: JR, VB, PWH, HK, LD, MK. Final approval of manuscript: JR, VB, PWH, HK, LD, MK. Accountable for all aspects of the work: JR, VB, PWH, HK, LD, MK.

## Abstract

**Background:** Even though fatigue is one of the most prevalent and burdensome symptoms in patients with glioma, its etiology and determinants are still poorly understood. We aimed to identify which demographic, tumor- and treatment-related characteristics and patient-reported outcome measures (PROMs) are associated with or are predictors of fatigue in glioma.

**Methods:** In this retrospective observational study, we included glioma patients with preoperative and postoperative assessments including PROMs on fatigue, depression, cognitive functioning, and health-related quality of life (HRQoL). Linear mixed models were used to identify which clinical factors and PROMs were associated with fatigue and linear multiple regression was used to detect predictors of postoperative fatigue.

**Results:** In this study, 222 patients were included (78% grade II-III glioma, 22% grade IV). These patients had performed 333 assessments (193 preoperative and 116 one year postoperatively). Of all assessments, 39% was indicative of severe fatigue. Several HRQoL domains, depression, and right-sided tumors were significantly associated with fatigue (marginal *R*^*2*^ = 0.63). Contrary to common expectations, tumor type, treatment-related factors and timing of the assessment, were not associated with fatigue. In a subgroup of seventy patients with follow-up assessments, preoperative fatigue and physical functioning were predictors of postoperative fatigue (adjusted *R*^*2*^ = 0.31).

**Conclusion:** Fatigue is a complex symptom, which should not solely be attributed to the tumor or its treatment, but is instead related to different aspects of mood and HRQoL. These insights are of importance in understanding fatigue and could guide symptom management, especially in patients with lower grade tumors.

**Key points:** Up to 40% of patients are severely fatigued, regardless of tumor type or timing of assessment HRQoL, depression, and right-sided tumor explain 63% of variance in fatigue severity Demographics, tumor type and treatment are not associated with fatigue severity

**Importance of the Study:** Despite fatigue being one of the most frequent and burdensome symptoms in patients with glioma, its etiology remains poorly understood. We aimed to unravel the impact of demographic, tumor- and treatment-related factors, symptoms, and HRQoL to grasp the complexity of fatigue in 222 glioma patients. Our findings demonstrate that fatigue is associated with several domains of HRQoL and depression, as well as having a tumor in the right hemisphere. What stands out is that fatigue is not associated with tumor type, treatment-related characteristics or phase of the disease. These findings could be important in symptom management since tumor- and treatment-related factors are non-modifiable, whilst depressive symptoms and social and physical functioning could be more suitable treatment targets. These results underscore the need for timely screening and symptom management focusing not only on fatigue but also on mood and diminished HRQoL.

## Background

Patients with diffuse glioma experience a multitude of symptoms during the course of the disease including fatigue, cognitive impairment, and neurological deficits.^1^ Fatigue is one of the most frequently reported and burdensome symptoms and is associated with a significant loss of health-related quality of life (HRQoL).^2^ The prevalence of fatigue fluctuates over time and reported numbers vary between studies.^3^ Of the lower-grade glioma patients 39-77% report fatigue,^4^ while 48% of glioblastoma patients report fatigue after surgery, and this number increases to up to 90% in patients with tumor recurrence.^5,6^ Because gliomas are still incurable and many patients suffer from fatigue and its sequelae, it is important to understand the interaction between fatigue and other symptoms and HRQoL to guide rational symptom management.

The etiology of fatigue in brain tumor patients is complex and poorly understood. Demographic, biomedical, neuropsychological, and psychosocial factors are thought to be related to fatigue.^7^ It is unclear whether those factors are causally linked to the emergence and perpetuation of fatigue, or that symptom burden and diminished HRQoL are a result of severe fatigue. Several studies have reported conflicting variables to be associated with fatigue in glioma patients, often in small samples including different types of primary brain tumor patients at different time points during the disease. A relatively large study identified poor performance status, female sex, and active disease (compared to stable disease) to be associated with fatigue in two-hundred primary brain tumor patients.^8^ Contrarily, another study in 65 postoperative glioblastoma patients found that performance status was not associated with fatigue, but that depression played an important role.^5^ Furthermore, two smaller studies in lower-grade glioma patients found that fatigued patients experienced more cognitive impairment, were older and used more anti-epileptic drugs.^9,10^ It is important to note, that all these studies were cross-sectional and did not take longitudinal changes of fatigue into account, and thus did not investigate any potential predictors of fatigue.

Fatigue is also a prominent symptom in patients with cancer, cancer survivors, and patients with neurological diseases, including multiple sclerosis and Parkinson’s disease.^11,12^ A meta-analysis combined fifteen studies on fatigue in common chronic disorders and found that over half of all patients suffered from severe fatigue and that type of disease could only explain a small proportion of variance in fatigue severity.^13^ Interestingly, when sex, age, motivational and concentration problems, pain, sleep disturbances, physical functioning, reduced activity and lower self-efficacy were added to the model, up to half of the variance in fatigue severity could be explained. The common denominator in those studies is that disease-specific factors were not associated with fatigue, whilst psycho-social symptoms, physical functioning and HRQoL were. It is not clear whether this would be the same in patients with glioma and whether tumor- and treatment-related factors play a role in glioma-related fatigue.

The current study aims to evaluate fatigue in a large sample of glioma patients in relation to demographic, tumor- and treatment-related variables, as well as symptoms and HRQoL measured with patient-reported outcome measures (PROMs). Furthermore, in a subset of patients with follow-up data, we aim to find predictors of postoperative fatigue and demonstrate how fatigue severity changes over time within the patient.

## Methods

### Participants and procedures

This is a retrospective observational study conducted at the Amsterdam UMC location Vrije Universiteit Amsterdam, a tertiary referral hospital in the Netherlands, consisting of a convenience sample of patients diagnosed with diffuse glioma. Ethical approval for the use of clinical data for research purposes was granted by the Medical Ethics Review Committee (METc VUmc 2010.126) and written informed consent was obtained from the patients.

Between June 2010 and April 2021, patients who were scheduled for elective tumor surgery were referred for assessment to the Department of Medical Psychology by their treating physician as part of routine clinical care. Both before and one year after surgery, patients conducted an assessment consisting of questionnaires on fatigue, depression, self-perceived cognitive functioning, brain tumor-specific symptoms, and HRQoL. If a patient was scheduled for re-resection, again assessments were conducted before and one year after re-resection. For the current study, patients who had conducted at least one assessment were included in the so-called ***Glioma cohort***. Patients in the Glioma cohort conducted a varying number of assessments (between one and six) as the result of re-resection or loss to follow-up. Assessments with a missing fatigue questionnaire were excluded. Additionally, to assess fatigue longitudinally, a selection of patients in the *Glioma cohort* were included in the ***Longitudinal subgroup***. These patients had performed both an assessment 0 to 6 months before surgery and an assessment 6 to 24 months after that same surgery.

### Patient-reported outcome measures

*Fatigue* was assessed with the Checklist Individual Strength (CIS), a 20-item questionnaire answered on a seven-point Likert scale focusing on symptoms in the past two weeks.^14^ The questionnaire consists of four subscales: fatigue severity (CIS-fatigue), concentration problems, reduced motivation, and reduced activity level. A cut-off of ≥35 on the CIS-fatigue subscale was used to indicate severe fatigue. The questionnaire and cut-off score have been validated in cancer survivors.^14^ A change of ≥7 points on the CIS-fatigue subscale was defined as clinically relevant improvement or deterioration.^15^

*Depression* was assessed with the Center for Epidemiologic Studies Depression questionnaire (CES-D).^16^ The Medical Outcomes Study Cognitive Functioning Scale (MOS-cog) was used to measure *self-perceived cognitive functioning*.^17^ *Health-related quality of life* was assessed with the Medical Outcomes Study Short-Form Health Survey (SF-36).^18^ The SF-36 is organized into nine scales assessing physical functioning, bodily pain, role limitations due to physical health problems, role limitations due to personal or emotional problems, emotional well-being, social functioning, energy/vitality, general health perceptions, and changes in health. In this study, we did not include the energy/vitality subscale. *Brain tumor-specific symptoms* were assessed with the European Organization for Research and Treatment of Cancer brain tumor module (BN20) questionnaire.^19^ We only included the items future uncertainty, motor dysfunction, and seizures in the analyses, because the other items were already assessed with the other questionnaires. All questionnaires were recoded and scored according to the relevant scoring manuals. A higher score of the CIS, CES-D, and BN-20 reflects more symptoms or problems, and a lower score of the MOS-cog and the SF-36 reflects more disability and worse functioning.

### Statistical analysis

The scripts for the statistical analyses are available in an online repository on Github (https://github.com/multinetlab-amsterdam/projects/tree/master/fatigue_glioma_manuscript_2022). These scripts can be run with the provided mock data in the script. Analyses were conducted using Rstudio (version 4.0.3).^20^ Two-sided *p*-values < 0.05 were considered significant. First, we presented the percentages of the assessments and patients who were classified as severely fatigued based on the CIS-fatigue cut-off score. To clarify, in the *Glioma cohort* the presented percentages reflect the number of assessments indicative of severe fatigue, and not the number of patients since multiple assessments per patient were included. In the *Longitudinal subgroup*, the percentages do reflect the number of fatigued patients preoperatively and postoperatively.

Our primary aim was to identify which demographic, tumor- and, treatment-related characteristics and PROMs were associated with fatigue severity in the *Glioma cohort* with a linear mixed model. Patients were included as random intercepts, so we could control for the inclusion of multiple assessments per patient. We took three distinct steps: first, with univariate linear mixed models we evaluated associations between the CIS-fatigue score as a continuous variable and age, sex, education, KPS, location of the tumor, histology (oligodendroglioma, astrocytoma, oligoastrocytoma, glioblastoma), isocitrate dehydrogenase (IDH) status (IDH mutation versus wildtype), phase of the disease (defined as pre-surgery, during chemoradiation and/or chemotherapy, stable disease, or progressive disease), the timing of the assessment (preoperative or postoperative), dexamethasone use (yes or no), radiotherapy before the assessment, chemotherapy before the assessment, and all the PROMs. Secondly, all above mentioned variables that showed a statistically significant association with fatigue were included in a linear mixed model as independent variables. As a third and final step, we performed backward selection based on the so-called Akaike Information Criterion to find the best-fitting model. After this, the model consisted of only variables that are significantly associated with fatigue severity. Together these variables can explain a proportion of the variance in the level of fatigue, demonstrated with an *R*^*2*^ value.

Our secondary aim was to find predictors of postoperative fatigue. The within-subject data of the *Longitudinal subgroup* was used to compute a multiple linear regression model with the postoperative CIS-fatigue score as the dependent variable and the clinical characteristics and preoperative PROMs as potential predictors. We used the same three-step approach as described above. First, we evaluated univariate associations between the postoperative CIS-fatigue score and the individual variables. Subsequently, all variables that were associated with the postoperative CIS-fatigue score were used to compute a multiple linear regression model. After backward selection, the final model included only clinical characteristics and preoperative PROMs that were statistically significant predictors of postoperative fatigue severity. Additionally, to assess within-subject similarity between preoperative and postoperative CIS-fatigue scores in the *Longitudinal subgroup*, we computed a two-way random effects single measure intraclass correlation coefficient with absolute agreement (ICC 2,1).^21^ The 95% confidence interval of the ICC was classified as poor (<0.5), moderate (0.5 – 0.75) or good (>0.75).^22^

## Results

In this study, 222 patients with diffuse glioma were included in the *Glioma cohort* (Figure 1). They had performed 343 assessments, of which ten were excluded because the CIS questionnaire was missing. Of the 333 assessments, 193 were conducted preoperatively and 116 postoperatively. The mean age of the 222 patients in the *Glioma cohort* was 45 years and 57% were male. The majority of tumors were located in the left hemisphere (62%) and the frontal lobe (60%) and were classified as a WHO grade II or III (Table 1). Patients in the *Glioma cohort* performed between 1 and 6 assessments. Of the *Glioma cohort*, seventy patients were included in the *Longitudinal subgroup* (Figure 1, Table 1). Almost all of these patients had a WHO grade II or III tumor. See Table S1 for an overview of the PROMs in the *Glioma cohort* and the *Longitudinal subgroup*.

**Table 1.** Sociodemographic and clinical characteristics *Glioma cohort* and *Longitudinal subgroup*. * Histological diagnosis of these patients was based on the 2007 WHO classification of the central nervous system tumors, and could not be re-classified because molecular markers were not available^49^ Abbreviations: SD, standard deviation; n, number.

|  | **Glioma cohort**<br>**(N=222)** | **Longitudinal**<br>**subgroup (n=70)** |
| --- | --- | --- |
| **Age, mean (SD)** | 45.6 (13.3) | 41.0 (11.6) |
| **Sex, n (%)** |  |  |
| Male | 126 (56.8%) | 42 (60.0%) |
| Female | 96 (43.2%) | 28 (40.0%) |
| **Level of education (Verhage), n (%)** |  |  |
| Low (1-4) | 41 (18.5%) | 11 (15.7%) |
| Middle (5) | 75 (33.8%) | 26 (37.1%) |
| High (6-7) | 106 (47.7%) | 32 (45.7%) |
| Missing |  | 1 (1.4%) |
| **Tumor hemisphere, n (%)** |  |  |
| Left | 137 (61.7%) | 46 (65.7%) |
| Right | 81 (36.5%) | 23 (32.9%) |
| Both | 4 (1.8%) | 1 (1.4%) |
| **Tumor location, n (%)** |  |  |
| Frontal | 132 (59.5%) | 41 (58.6%) |
| Non-frontal | 90 (40.5%) | 29 (41.4%) |
| **WHO grade, n (%)** |  |  |
| II | 114 (51.4%) | 47 (67.1%) |
| III | 60 (27.0%) | 20 (28.6%) |
| IV | 48 (21.6%) | 3 (4.3%) |
| **Tumor type, n (%)** |  |  |
| Oligodendroglioma | 74 (33.3%) | 33 (47.1%) |
| Astrocytoma, IDH mutated | 67 (30.2%) | 24 (34.3%) |
| Astrocytoma, IDH wildtype | 10 (4.5%) | 3 (4.3%) |
| Astrocytoma, IDH status unknown | 14 (6.3%) | 3 (4.3%) |
| Oligoastrocytoma* | 9 (4.1%) | 4 (5.7%) |
| Glioblastoma, IDH mutated | 5 (2.3%) | 1 (1.4%) |
| Glioblastoma, IDH wildtype | 21 (9.5%) | 2 (2.9%) |
| Glioblastoma, IDH status unknown | 22 (9.9%) | 0 (0%) |
| **Using anti-epileptic drugs, n (%)** |  |  |
| No | 5 (2.3%) | 1 (1.4%) |
| Yes | 163 (73.4%) | 52 (74.3%) |
| Missing | 54 (24.3%) | 17 (24.3%) |
| **Number of included assessments per patient, n (%)** |  |  |
| 1 | 137 (61.7%) |  |
| 2 | 68 (30.3%) | 70 (100%) |
| 3 | 10 (4.5%) |  |
| 4 | 6 (2.7%) |  |
| 6 | 1 (0.5%) |  |

**Figure 1.**
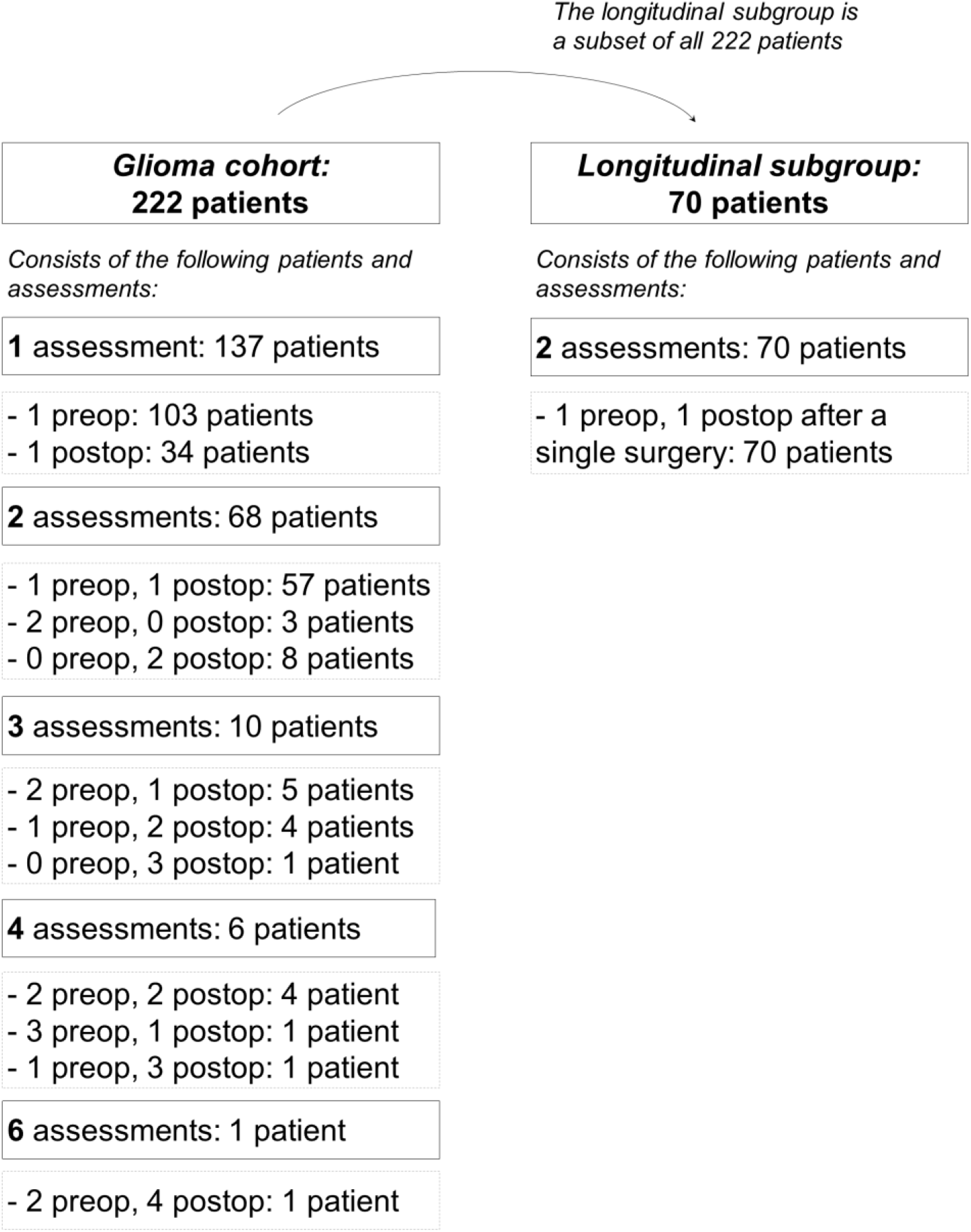
Included patients and assessments in *Glioma cohort* and the *Longitudinal subgroup*. Abbreviations: Preop, preoperative assessment; Postop, postoperative assessment

### Prevalence of fatigue in the Glioma cohort and the Longitudinal subgroup

Of all assessments in the *Glioma cohort*, 39% (130/333) were indicative of severe fatigue (Figure 2). Of the preoperative assessments 38% were classified as severely fatigued, and 41% of the postoperative assessments. During treatment, 37% of the assessments (30/81) were indicative of severe fatigue, and 43% (33/76) of the assessments in patients with stable disease and 38% (9/24) of the assessments in patients with progressive disease. In the *Longitudinal subgroup* preoperatively 34% of the patients and postoperatively 40% of the patients were severely fatigued.

**Figure 2.**
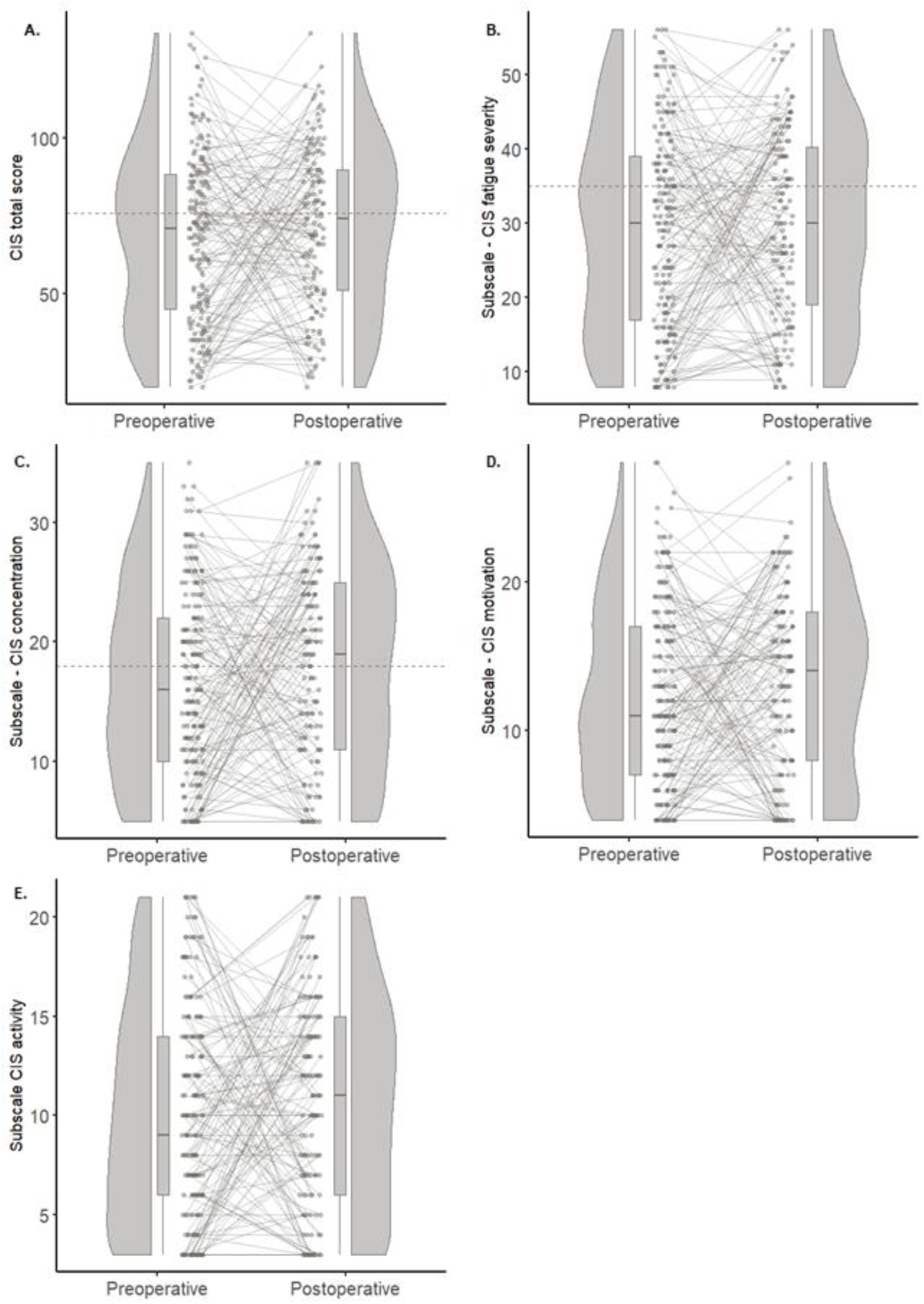
Raincloud plots preoperative and postoperative Checklist Individual Strength. Each panel shows a raincloud plot, which is a combination of a scatterplot, a boxplot with median and interquartile range, and half a violin plot ^50^. The grey lines between the dots of the scatterplots are combinations of scores of a preoperative and postoperative assessment within one patient. The grey dotted horizontal lines indicate the cut-off scores for the scales with available validated cut-off values. All panels include all assessments of the Glioma cohort. Panel A reflects the Checklist Individual Strength (CIS) total score. Panels B-E reflect the four CIS subscales. Abbreviations: CIS, checklist individual strength.

### Explaining variance in fatigue severity in the Glioma cohort

In the *Glioma cohort*, depressive symptoms, four HRQoL domains, and having a tumor in the right hemisphere were significantly associated with fatigue severity (CIS-fatigue score), and explained a large proportion of variance in fatigue severity between patients. First, with univariate comparisons, we determined that age, dexamethasone use, chemotherapy, radiotherapy, tumor grade, histology, involved lobes, IDH status, phase of the disease, and timing of the assessment were not associated with fatigue severity (Table 2). On the other hand, depressive symptoms, self-perceived cognitive functioning, brain-tumor related symptoms, all eight HRQoL domains, sex, education, tumor hemisphere, and KPS were independently associated with fatigue severity. These significantly associated variables were included in a linear mixed model, and after backward selection, the final model included depressive symptoms, four HRQoL domains and having a tumor in the right hemisphere (Table 3). These six variables were significantly associated with fatigue severity and together explained 63% of the variance in fatigue severity (marginal *R*^*2*^ = 0.63).

**Table 2.**
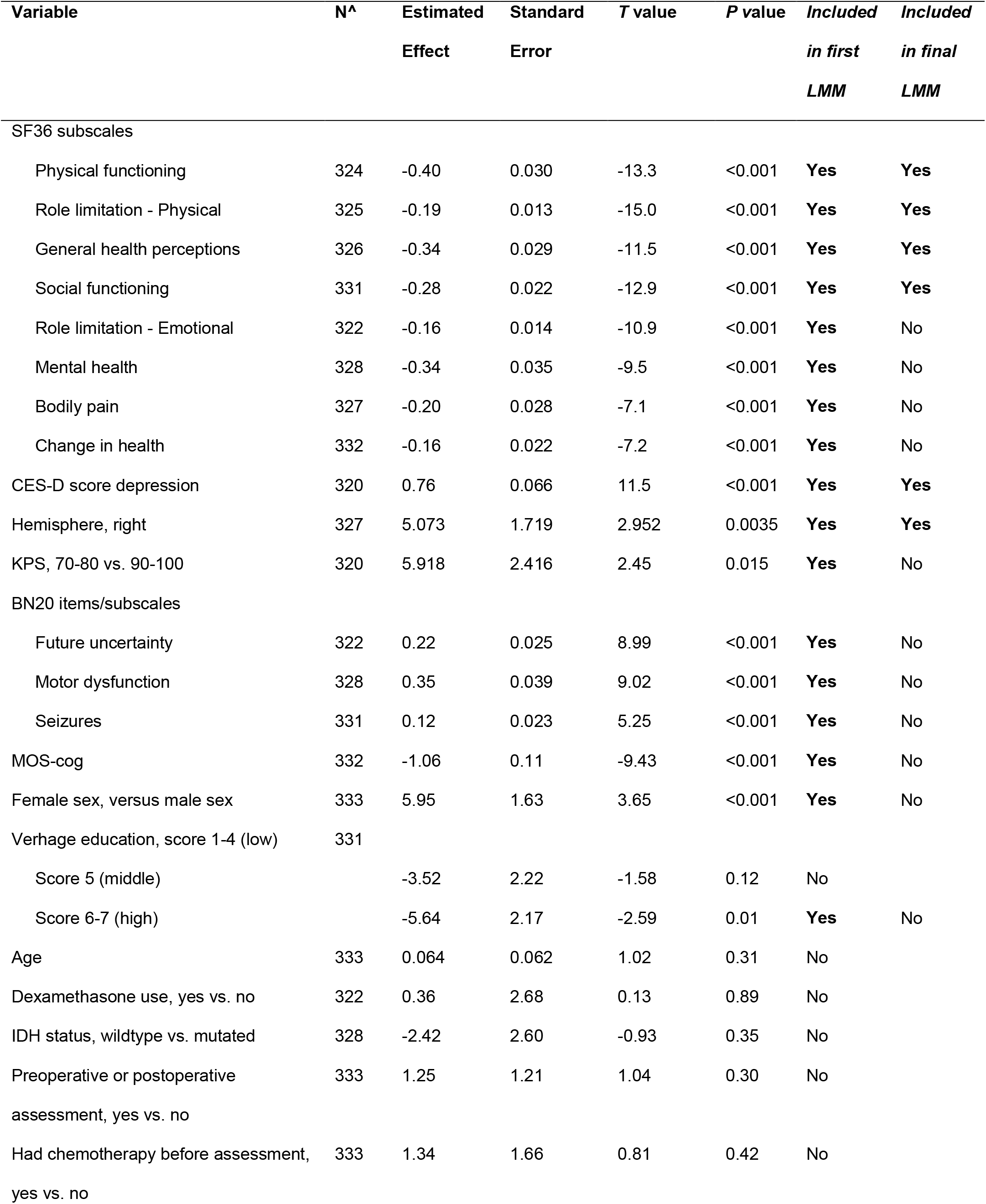
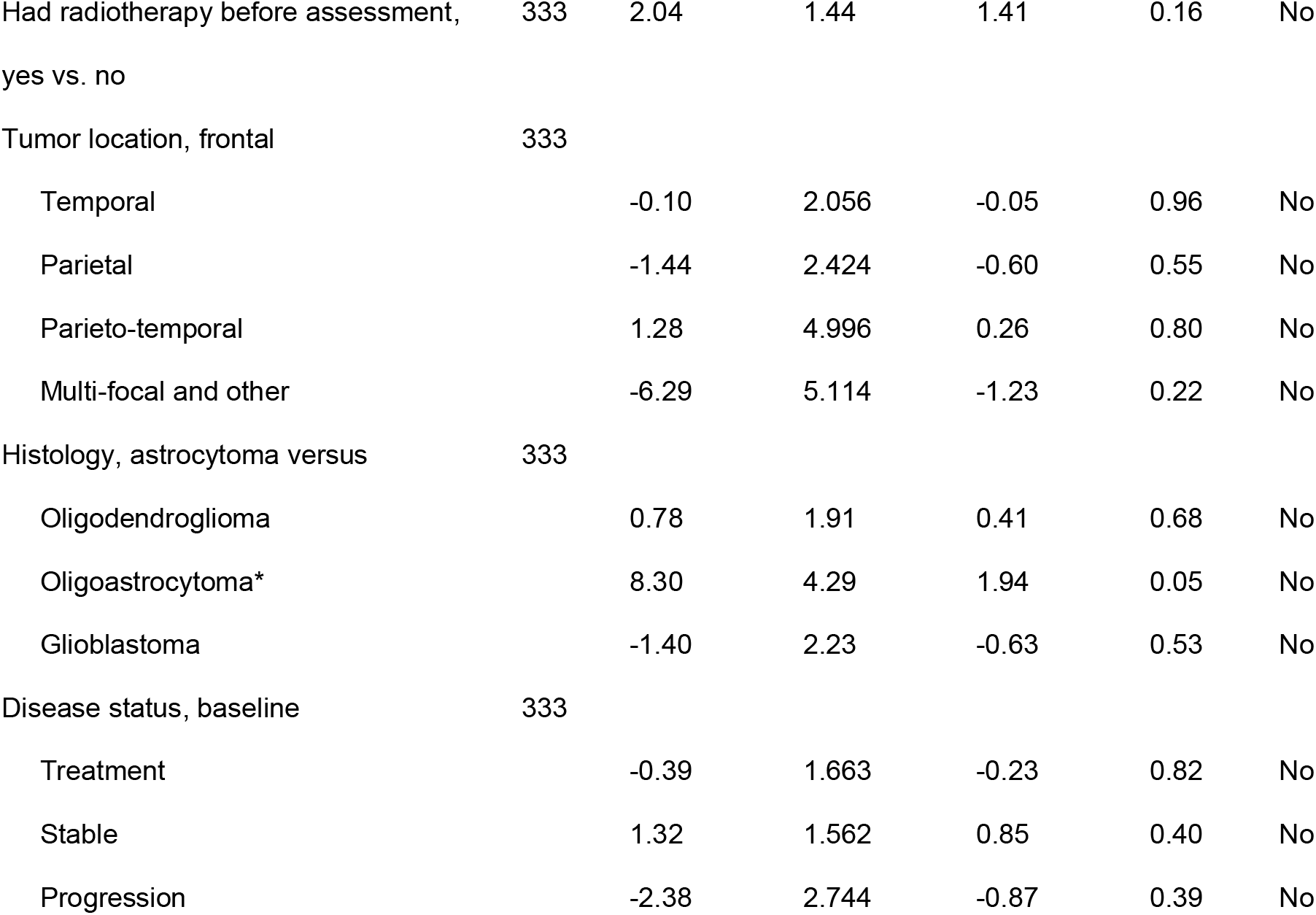
Associations between CIS-fatigue subscale and clinical factors and PROMs. For each variable, a linear mixed model was computed with CIS-fatigue as the dependent factor and patient as a random effect. For example, the SF-36 physical functioning subscale was significantly associated with the CIS-fatigue score. This variable was then included in the next step of the analysis and after backward selection remained included in the final linear mixed model. ^ N is the number of assessments included in the analysis * Histological diagnosis of these patients was based on the 2007 WHO classification of the central nervous system tumors, and could not be re-classified because molecular markers were not available^49^ Abbreviations: CES-D, Center for Epidemiologic Studies Depression; CIS-fatigue, Checklist Individual Strength subscale fatigue severity; IDH, Isocitrate dehydrogenase; MOS-cog, Medical Outcomes Study Cognitive Functioning Scale; SF36, Medical Outcomes Study Short-Form Health Survey; BN20, The European Organization for Research and Treatment of Cancer brain tumor module.

| Variable | N^ | Estimated<br>Effect | Standard<br>Error | T value | P value | Included<br>in first<br>LMM | Included<br>in final<br>LMM |
| --- | --- | --- | --- | --- | --- | --- | --- |
| SF36 subscales |  |  |  |  |  |  |  |
| Physical functioning | 324 | -0.40 | 0.030 | -13.3 | <0.001 | Yes | Yes |
| Role limitation - Physical | 325 | -0.19 | 0.013 | -15.0 | <0.001 | Yes | Yes |
| General health perceptions | 326 | -0.34 | 0.029 | -11.5 | <0.001 | Yes | Yes |
| Social functioning | 331 | -0.28 | 0.022 | -12.9 | <0.001 | Yes | Yes |
| Role limitation - Emotional | 322 | -0.16 | 0.014 | -10.9 | <0.001 | Yes | No |
| Mental health | 328 | -0.34 | 0.035 | -9.5 | <0.001 | Yes | No |
| Bodily pain | 327 | -0.20 | 0.028 | -7.1 | <0.001 | Yes | No |
| Change in health | 332 | -0.16 | 0.022 | -7.2 | <0.001 | Yes | No |
| CES-D score depression | 320 | 0.76 | 0.066 | 11.5 | <0.001 | Yes | Yes |
| Hemisphere, right | 327 | 5.073 | 1.719 | 2.952 | 0.0035 | Yes | Yes |
| KPS, 70-80 vs. 90-100 | 320 | 5.918 | 2.416 | 2.45 | 0.015 | Yes | No |
| BN20 items/subscales |  |  |  |  |  |  |  |
| Future uncertainty | 322 | 0.22 | 0.025 | 8.99 | <0.001 | Yes | No |
| Motor dysfunction | 328 | 0.35 | 0.039 | 9.02 | <0.001 | Yes | No |
| Seizures | 331 | 0.12 | 0.023 | 5.25 | <0.001 | Yes | No |
| MOS-cog | 332 | -1.06 | 0.11 | -9.43 | <0.001 | Yes | No |
| Female sex, versus male sex | 333 | 5.95 | 1.63 | 3.65 | <0.001 | Yes | No |
| Verhage education, score 1-4 (low) | 331 |  |  |  |  |  |  |
| Score 5 (middle) |  | -3.52 | 2.22 | -1.58 | 0.12 | No |  |
| Score 6-7 (high) |  | -5.64 | 2.17 | -2.59 | 0.01 | Yes | No |
| Age | 333 | 0.064 | 0.062 | 1.02 | 0.31 | No |  |
| Dexamethasone use, yes vs. no | 322 | 0.36 | 2.68 | 0.13 | 0.89 | No |  |
| IDH status, wildtype vs. mutated | 328 | -2.42 | 2.60 | -0.93 | 0.35 | No |  |
| Preoperative or postoperative<br>assessment, yes vs. no | 333 | 1.25 | 1.21 | 1.04 | 0.30 | No |  |
| Had chemotherapy before assessment,<br>yes vs. no | 333 | 1.34 | 1.66 | 0.81 | 0.42 | No |  |

**Table 2. Associations between CIS-fatigue subscale and clinical factors and PROMs**
|  |  |  |  |  |  |  |
| --- | --- | --- | --- | --- | --- | --- |
| Had radiotherapy before assessment, yes vs. no | 333 | 2.04 | 1.44 | 1.41 | 0.16 | No |
| Tumor location, frontal | 333 |  |  |  |  |  |
| Temporal |  | -0.10 | 2.056 | -0.05 | 0.96 | No |
| Parietal |  | -1.44 | 2.424 | -0.60 | 0.55 | No |
| Parieto-temporal |  | 1.28 | 4.996 | 0.26 | 0.80 | No |
| Multi-focal and other |  | -6.29 | 5.114 | -1.23 | 0.22 | No |
| Histology, astrocytoma versus | 333 |  |  |  |  |  |
| Oligodendroglioma |  | 0.78 | 1.91 | 0.41 | 0.68 | No |
| Oligoastrocytoma* |  | 8.30 | 4.29 | 1.94 | 0.05 | No |
| Glioblastoma |  | -1.40 | 2.23 | -0.63 | 0.53 | No |
| Disease status, baseline | 333 |  |  |  |  |  |
| Treatment |  | -0.39 | 1.663 | -0.23 | 0.82 | No |
| Stable |  | 1.32 | 1.562 | 0.85 | 0.40 | No |
| Progression |  | -2.38 | 2.744 | -0.87 | 0.39 | No |

**Table 3.** HRQoL, depression, lesion in the right hemisphere associated with fatigue severity. This table shows the best-fit linear mixed model with six variables explaining a large proportion of variance in the CIS-fatigue score. Patient was included as a random effect to control for multiple measurements. Abbreviations: CES-D, Center for Epidemiologic Studies Depression; CIS-fatigue, Checklist Individual Strength subscale fatigue severity; SF-36, Medical Outcomes Study Short-Form Health Survey.

| Fixed effects | Estimated effect | Standard error | T value | P value |
| --- | --- | --- | --- | --- |
| Intercept: CIS-fatigue | 53.568 | 3.181 | 16.8 | <0.001 |
| SF-36 role limitation - physical | -0.086 | 0.015 | -5.8 | <0.001 |
| SF-36 physical functioning | -0.169 | 0.032 | -5.2 | <0.001 |
| SF-36 general health perceptions | -0.121 | 0.028 | -4.3 | <0.001 |
| SF-36 social functioning | -0.054 | 0.026 | -2.1 | 0.039 |
| CES-D | 0.234 | 0.069 | 3.4 | <0.001 |
| Right hemisphere | 3.353 | 1.096 | 3.1 | 0.003 |

### Within-subject changes and predictors of postoperative fatigue

In the *Longitudinal subgroup*, fatigue improved postoperatively in 24% of the patients (17/70) and worsened in 30% (21/70, Figure 3). Within-patient similarity between the preoperative and postoperative fatigue severity was poor to moderate (ICC = 0.522, 95% CI 0.329 - 0.673), indicating that the level of fatigue indeed often fluctuated within the patient.

**Figure 3.**
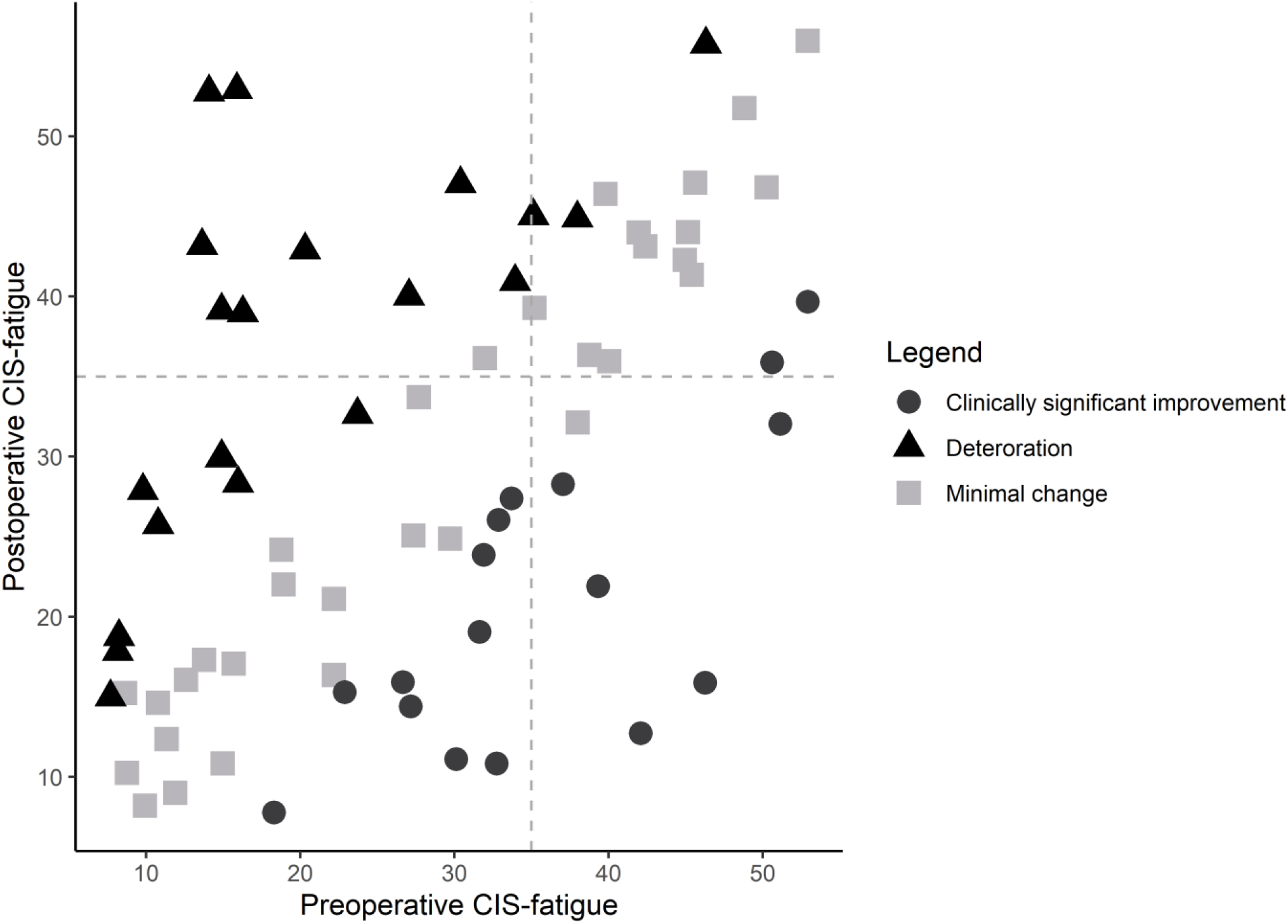
Improvement and deterioration of CIS-fatigue scores in the *Longitudinal subgroup*. The horizontal and vertical grey dashed lines reflect the cut-off score of the CIS-fatigue of 35 points. Deteriorating is defined as an increase in CIS-fatigue of ≥ 7, clinically significant improvement as a decrease in CIS-fatigue of ≥ 7.^15^

Moreover, when patients experience high levels of fatigue preoperatively or lowered physical functioning (SF36-PF) preoperatively, these patients are more likely to experience high levels of fatigue one year postoperatively. First, with univariate comparisons, we found that demographic and tumor- and treatment-related variables were not predictive of postoperative fatigue severity (Table S2), but that the preoperative fatigue subscales, depressive symptoms, self-perceived cognitive functioning, brain-tumor related symptoms, and HRQoL domains were significant predictors of postoperative fatigue severity. After combining these significantly associated variables in one multiple linear regression model and applying backward selection, the final model identified preoperative fatigue severity (estimated effect 0.32, *p =* 0.02) and preoperative physical functioning (estimated effect -0.26, *p =* 0.02) as significant predictors of postoperative fatigue (adjusted *R*^*2*^ = 0.31).

## Discussion

The current study aimed to identify which demographic, tumor- and treatment-related characteristics and PROMs were associated with fatigue severity in a large sample of glioma patients and which factors were predictive of postoperative fatigue in a subgroup of patients with both preoperative and postoperative assessments. We showed that a large proportion of the variance in fatigue severity could be explained by patient-reported limitations in HRQoL, depressive symptoms, and a right-sided tumor. Preoperative fatigue and physical functioning were independent predictors of postoperative fatigue.

Interestingly, there were no direct associations between fatigue severity and age, tumor type, treatment characteristics, phase of the disease, or timing of the assessment. These results corroborate studies in brain tumor patients that found no link between fatigue and disease duration, type of neurosurgical intervention, and chemo- and radiotherapy.^8-10^ Also in cancer survivors and patients with chronic disease, the persistence of fatigue is not determined by tumor- and treatment-related characteristics.^11,23-25^ More importantly, sleep disturbances, reduced physical functioning and activity, and lower self-efficacy concerning fatigue, do seem to explain the differences between fatigued and non-fatigued patients.^11,13,26^ In line with these studies, we found limitations in HRQoL and depressive symptoms to be associated with fatigue, rather than tumor- and treatment-related factors. Functional and physical impairments have been identified as important determinants of fatigue and HRQoL before.^8,27,28^ Similarly, depression is also known to be associated with fatigue in patients with cancer, meningioma, and neurological disease, with generally high correlations between the two.^29-31^ However, it does remain to be elucidated whether fatigue is a direct result of depression and diminished HRQoL, or whether the association between the two is bidirectional or even reversed. Future studies should focus on why fatigue persists or emerges in some patients, while it decreases in others, for example through the incorporation of patients’ daily life reports in research using ecological momentary assessments.^32^

We found tumors in the right hemisphere to be associated with elevated levels of fatigue. Tumor laterality has inconsistently been shown to be associated with HRQoL and symptoms like fatigue, depression and anxiety.^33-35^ Also in stroke patients, there is no conclusive evidence on the association between fatigue and depression, and hemisphere and lesion location.^36-38^ A recent study showed right-hemisphere gliomas to be associated with worse HRQoL, compared to left-hemisphere tumors.^39^ The authors hypothesized that tumor infiltration in areas involved with controlling empathy could result in higher self-focus with diminished HRQoL as an effect. Several models explain the possible hemispheric lateralization of emotions, for example how primary emotions are controlled by the right hemisphere and social emotion by the left hemisphere.^40^ Perhaps these proposed mechanisms could also explain the association between fatigue and tumor laterality.

In the *Longitudinal subgroup*, about 40% of the patients reported severe fatigue preoperatively, and half of the patients showed clinical improvement or deterioration of the CIS-fatigue score. We identified preoperative fatigue and physical functioning as independent predictors of postoperative fatigue. Fatigue at baseline has been identified as an important predictor of fatigue at follow-up in cancer patients before.^41^ In line with our study, research in women with breast cancer found that preoperative fatigue and lower role functioning before surgery, and not cancer characteristics, were associated with heightened postoperative fatigue levels.^42^ Clinically, these results underline that preoperative identification of glioma patients suffering from fatigue or impaired physical functioning is important. Potentially, preoperative fatigue treatment and symptom management could also result in less symptom burden later on. However, this remains to be investigated with intervention studies with adequate follow-up.

Throughout this study, we have made a distinction between PROMs and demographic, and tumor- and treatment-related factors. This distinction is of importance in symptom management and when developing interventions targeting fatigue. Even though tumor- and treatment-related factors are important in disease management, these factors are usually non-modifiable and therefore not suitable as targets in ameliorating fatigue.^43^ On the other hand, factors such as depression, social and physical functioning and health perception are modifiable to some extent and therefore could be relevant targets. Moreover, these results call for the timely and integrative assessment and treatment of fatigue, psychological symptoms and impaired HRQoL in glioma patients. Additionally, it could be of interest to understand whether mastery, or sense of control, coping style and catastrophizing are associated with fatigue in glioma patients. It is already known, that among patients with cancer a high sense of personal mastery is associated with less fatigue.^44^ Also, decreased catastrophizing is a mediator in trials investigating the effectiveness of cognitive-behavioral therapy targeting fatigue. Currently, there are no evidence-based interventions targeting fatigue in glioma patients.^7^ However, there are several effective interventions targeting fatigue in cancer patients, including cognitive behavioral therapy, mindfulness, exercise, activity enhancement, problem-solving therapy, and acceptance and commitment therapy.^24,45^ Those interventions might also be promising in treating fatigue in patients with glioma.^45-47^ Unfortunately, we did not assess whether antiepileptic drugs (AEDs) were associated with fatigue, since some studies do suggest that AEDs could result in fatigue.^5,10,48^ Therefore it might be worthwhile to investigate whether switching between AEDs or lowering the dosage would be a useful approach to ameliorate fatigue.^48^

An important limitation of this study is the potential selection bias introduced by the procedure of active referral. Patients included in this study were referred to the Department of Medical Psychology by their treating physician, and therefore this study consists of a convenience sample. It is likely that physicians more often referred younger patients, patients with tumors in eloquent areas, or patients scheduled for awake craniotomy. Furthermore, caution is warranted since not all patients participated in follow-up assessments. Almost all of the patients included in the *Longitudinal subgroup* had a WHO grade II or III tumor, hence these longitudinal results are mainly applicable to patients with a lower grade tumor. Additionally, we acknowledge that a wide range of questionnaires and PROMs are available to address symptoms, all with varying psychometric properties. With regards to the CIS questionnaire, it is unknown how changes in the CIS score during a follow-up period reflect actual changes in everyday life in the individual patient. The CIS questionnaire does have high test-retest reliability (r=0.74-0.86) but unfortunately has only been validated in cancer survivors, and not in glioma patients.^14^ In the Dutch population,15% of the patients without chronic disease report severe fatigue based on the CIS-fatigue score,^11^ compared to around 40% in the current study, indicating the prevalence of fatigue is indeed much higher in glioma patients. However, it would be of value to replicate the current results using other questionnaires and PROMs.

In conclusion, fatigue is a highly prevalent symptom throughout the disease in patients with glioma. Tumor type, treatment-related factors, the timing of the assessment, and phase of the disease were not associated with fatigue severity, whilst physical functioning, physical role limitations, general health perceptions, social functioning, depressive symptoms, and a right-sided tumor were. In lower grade glioma patients, levels of fatigue vary substantially over time, and preoperative fatigue and physical functioning are important predictors of fatigue one year after surgery. The current study demonstrates that fatigue is a complex symptom, that should not solely be contributed to the tumor itself or its treatment but is related to a broad range of symptoms. Since the current study included patients with predominantly lower grade tumors, we recommend corroborating the current findings in a sample with more glioblastoma patients with an extensive longitudinal approach. In clinical care, the interplay between fatigue, depression, and HRQoL factors, should be taken into account when providing care for fatigued glioma patients and when designing interventions to target fatigue.

## Data Availability

All data produced in the present study are available upon reasonable request to the authors. All scripts for the analyses are are available online at Github (https://github.com/multinetlab-amsterdam/projects/tree/master/fatigue_glioma_manuscript_2022). These scripts can be run with the provided mock data in the script.

## Abbreviations

AEDs: Antiepileptic drugs
BN20: The European Organization for Research and Treatment of Cancer brain tumor module
CES-D: Center for Epidemiologic Studies Depression
CIS: Checklist Individual Strength
CIS-fatigue: Checklist Individual Strength subscale fatigue severity
HRQoL: Health-related quality of life
ICC: Intraclass correlation coefficient
IDH: Isocitrate dehydrogenase
MOS-cog: Medical Outcomes Study Cognitive Functioning Scale
PROM: Patient-reported outcome measure
SF-36: Medical Outcomes Study Short-Form Health Survey

