## Supplementary for "Toward unraveling the correlates of fatigue in glioma"

### Supplementary materials

***Table S1. Outcome of the questionnaires***

All values are presented as median [interquartile range]. Missing values are presented as number (percentage).

| | Preoperative assessments (N=193) | Postoperative assessments (N=140) | All assessments (N=333) |
|---|---|---|---|
| CIS total score | 71.0 [45.0, 88.5] | 74.0 [51.0, 91.5] | 72.0 [47.0, 90.0] |
| Missing | 2 (1.0%) | 2 (1.4%) | 4 (1.2%) |
| CIS subjective fatigue | 30.0 [17.0, 39.0] | 30.0 [19.0, 40.3] | 30.0 [18.0, 40.0] |
| Missing | 0 (0%) | 0 (0%) | 0 (0%) |
| CIS concentration | 16.0 [10.0, 22.0] | 19.0 [11.0, 25.0] | 17.0 [11.0, 24.0] |
| Missing | 2 (1.0%) | 0 (0%) | 2 (0.6%) |
| CIS motivation | 11.0 [7.00, 17.0] | 14.0 [8.00, 18.0] | 12.0 [7.00, 17.0] |
| Missing | 0 (0%) | 1 (0.7%) | 1 (0.3%) |
| CIS activity | 9.00 [6.00, 14.0] | 11.0 [6.00, 15.0] | 10.0 [6.00, 14.0] |
| Missing | 0 (0%) | 1 (0.7%) | 1 (0.3%) |
| MOS-cog | 29.0 [25.0, 33.0] | 27.0 [22.0, 31.5] | 28.0 [24.0, 33.0] |
| Missing | 0 (0%) | 1 (0.7%) | 1 (0.3%) |
| CES-D score depression | 10.0 [5.00, 16.8] | 11.0 [4.25, 18.0] | 10.5 [5.00, 17.3] |
| Missing | 3 (1.6%) | 10 (7.1%) | 13 (3.9%) |
| SF-36 Physical functioning | 90.0 [80.0, 100] | 90.0 [65.0, 100] | 90.0 [70.0, 100] |
| Missing | 4 (2.1%) | 5 (3.6%) | 9 (2.7%) |
| SF-36 Social functioning | 75.0 [50.0, 87.5] | 62.5 [50.0, 96.9] | 75.0 [50.0, 87.5] |
| Missing | 0 (0%) | 2 (1.4%) | 2 (0.6%) |
| SF-36 Role limitation - Physical | 25.0 [0, 100] | 25.0 [0, 100] | 25.0 [0, 100] |
| Missing | 3 (1.6%) | 5 (3.6%) | 8 (2.4%) |
| SF-36 Role limitation - Emotional | 100 [33.3, 100] | 66.7 [0, 100] | 66.7 [0, 100] |
| Missing | 4 (2.1%) | 7 (5.0%) | 11 (3.3%) |
| SF-36 Mental Health | 72.0 [56.0, 84.0] | 72.0 [56.0, 84.0] | 72.0 [56.0, 84.0] |
| Missing | 3 (1.6%) | 2 (1.4%) | 5 (1.5%) |
| SF-36 Energy / Vitality | 60.0 [45.0, 75.0] | 55.0 [40.0, 70.0] | 60.0 [45.0, 75.0] |
| Missing | 0 (0%) | 4 (2.9%) | 4 (1.2%) |
| SF-36 Bodily pain | 89.8 [67.3, 100] | 87.8 [67.3, 100] | 89.8 [67.3, 100] |
| Missing | 0 (0%) | 6 (4.3%) | 6 (1.8%) |
| SF-36 General health perceptions | 55.0 [40.0, 70.0] | 55.0 [40.0, 70.0] | 55.0 [40.0, 70.0] |
| Missing | 5 (2.6%) | 2 (1.4%) | 7 (2.1%) |
| SF-36 Change in health | 50.0 [25.0, 50.0] | 50.0 [25.0, 75.0] | 50.0 [25.0, 50.0] |
| Missing | 1 (0.5%) | 0 (0%) | 1 (0.3%) |
| BN20 future uncertainty | 41.7 [25.0, 66.7] | 33.3 [16.7, 58.3] | 41.7 [25.0, 58.3] |

| | Preoperative assessments (N=193) | Postoperative assessments (N=140) | All assessments (N=333) |
|---|---|---|---|
| Missing | 5 (2.6%) | 6 (4.3%) | 11 (3.3%) |
| BN20 visual disorder | 0 [0, 22.2] | 0 [0, 22.2] | 0 [0, 22.2] |
| Missing | 1 (0.5%) | 2 (1.4%) | 3 (0.9%) |
| BN20 motor dysfunction | 0 [0, 22.2] | 11.1 [0, 22.2] | 11.1 [0, 22.2] |
| Missing | 1 (0.5%) | 4 (2.9%) | 5 (1.5%) |
| BN20 communication deficit | 11.1 [0, 33.3] | 22.2 [2.78, 33.3] | 22.2 [0, 33.3] |
| Missing | 1 (0.5%) | 2 (1.4%) | 3 (0.9%) |
| BN20 headaches | 33.3 [0, 33.3] | 33.3 [0, 33.3] | 33.3 [0, 33.3] |
| Missing | 0 (0%) | 1 (0.7%) | 1 (0.3%) |
| BN20 seizures | 0 [0, 33.3] | 0 [0, 0] | 0 [0, 33.3] |
| Missing | 1 (0.5%) | 1 (0.7%) | 2 (0.6%) |
| BN20 drowsiness | 33.3 [0, 33.3] | 33.3 [0, 33.3] | 33.3 [0, 33.3] |
| Missing | 0 (0%) | 1 (0.7%) | 1 (0.3%) |
| BN20 hair loss | 0 [0, 0] | 0 [0, 0] | 0 [0, 0] |
| Missing | 0 (0%) | 1 (0.7%) | 1 (0.3%) |
| BN20 itchy skin | 0 [0, 0] | 0 [0, 33.3] | 0 [0, 33.3] |
| Missing | 0 (0%) | 1 (0.7%) | 1 (0.3%) |
| BN20 weakness of legs | 0 [0, 0] | 0 [0, 0] | 0 [0, 0] |
| Missing | 0 (0%) | 1 (0.7%) | 1 (0.3%) |
| BN20 bladder control | 0 [0, 0] | 0 [0, 0] | 0 [0, 0] |
| Missing | 1 (0.5%) | 2 (1.4%) | 3 (0.9%) |

*Table S2. Univariate comparisons between CIS-fatigue and potential predictor variables*

| Variable | N | Estimated Effect | Standard Error | *t*-value | *p*-value | *Included in first MLRM** | *Included in final MLRM^* |
|---|---|---|---|---|---|---|---|
| **Preoperative questionnaires** | | | | | | | |
| Checklist individual strength (CIS) | | | | | | | |
| CIS-fatigue | 70 | 0.521 | 0.103 | 5.047 | <0.001 | **Yes** | **Yes** |
| CIS-concentration | 70 | 0.615 | 0.187 | 3.292 | 0.002 | **Yes** | No |
| CIS-motivation | 70 | 0.914 | 0.247 | 3.697 | <0.001 | **Yes** | No |
| CIS-activity | | 1.150 | 0.254 | 4.529 | <0.001 | **Yes** | No |
| SF-36 subscales | | | | | | | |
| Physical functioning | 70 | -0.436 | 0.087 | -4.994 | <0.001 | **Yes** | **Yes** |
| Social functioning | 70 | -0.202 | 0.060 | -3.391 | 0.001 | **Yes** | No |
| Role limitation - Physical | 69 | -0.140 | 0.036 | -3.885 | <0.001 | **Yes** | No |
| Role limitation - Emotional | 70 | -0.081 | 0.039 | -2.099 | 0.040 | **Yes** | No |
| Mental health | 70 | -0.187 | 0.086 | -2.179 | 0.033 | **Yes** | No |
| Bodily pain | 70 | -0.146 | 0.066 | -2.21 | 0.031 | **Yes** | No |
| General health perceptions | 68 | -0.248 | 0.069 | -3.567 | <0.001 | **Yes** | No |
| Change in health | 69 | -0.149 | 0.072 | -2.067 | 0.043 | **Yes** | No |
| MOS-cog | 70 | -0.826 | 0.285 | -2.895 | 0.005 | **Yes** | No |
| CES-D score depression | 69 | 0.586 | 0.155 | 3.775 | <0.001 | **Yes** | No |
| BN20 items/subscales | | | | | | | |
| Future uncertainty | 69 | 0.182 | 0.063 | 2.918 | 0.005 | **Yes** | No |
| Motor dysfunction | 69 | 0.243 | 0.102 | 2.380 | 0.020 | **Yes** | No |
| Seizures | 70 | 0.057 | 0.053 | 1.065 | 0.291 | No | |
| **Demographic variables** | | | | | | | |
| Female sex | 70 | 5.726 | 3.289 | 1.741 | 0.086 | No | No |
| Age | 70 | 0.043 | 0.143 | 0.300 | 0.765 | No | No |
| Verhage education, score 1-4 (low) | 69 | | | | | | |
| Score 5 (middle) | | -8.647 | 4.862 | -1.778 | 0.080 | No | No |
| Score 6-7 (high) | | -8.330 | 4.725 | -1.763 | 0.083 | No | No |
| **Tumor and treatment characteristics** | | | | | | | |
| IDH status | 68 | -9.947 | 5.976 | -1.665 | 0.101 | No | No |
| Right hemisphere | 69 | 3.717 | 3.516 | 1.057 | 0.294 | No | No |
| Tumor location, frontal | | | | | | | |
| Temporal | 70 | -3.279 | 4.003 | -0.819 | 0.416 | No | No |
| Parietal | 70 | -1.462 | 4.318 | -0.338 | 0.736 | No | No |
| Histology, astrocytoma versus | 70 | | | | | | |
| Oligodendroglioma | | -0.988 | 3.520 | -0.281 | 0.780 | No | No |
| Oligoastrocytoma | | 2.883 | 7.427 | 0.388 | 0.699 | No | No |
| Glioblastoma | | -0.867 | 8.449 | -0.103 | 0.919 | No | No |
| Dexamethasone use *preoperative* | 70 | -1.530 | 8.186 | -0.187 | 0.852 | No | No |
| Karnofsky performance score *preoperative* | 68 | -7.941 | 5.810 | -1.367 | 0.176 | No | No |
| Had chemotherapy before postoperative | 70 | -0.821 | 3.360 | -0.244 | 0.808 | No | No |
| Had radiotherapy before postoperative | 70 | -1.583 | 3.322 | -0.477 | 0.635 | No | No |

* Variables that were included in the multiple linear regression model before backward selection
^ Variables that were included in the final multiple linear regression model after backward selection

*Abbreviations:*

*CES-D, Center for Epidemiologic Studies Depression; IDH, Isocitrate dehydrogenase; MOS-cog, Medical Outcomes Study Cognitive Functioning Scale; N, number; SF-36, Medical Outcomes Study Short-Form Health Survey; BN20, The European Organization for Research and Treatment of Cancer brain tumor module.*
